# Analytic validation of an *FGFR*-focused cell-free DNA liquid biopsy assay (FGFR-Dx)

**DOI:** 10.1101/2024.09.01.24312783

**Authors:** Julie W. Reeser, Michele R. Wing, Eric Samorodnitsky, Thuy Dao, Amy Smith, Leah Stein, Anoosha Paruchuri, Jharna Miya, Russell Bonneville, Yi-Seok Chang, Matthew Avenarius, Aharon G. Freud, Lianbo Yu, Sameek Roychowdhury

## Abstract

Commercial liquid biopsy assays are routinely used by oncologists to monitor disease response and resistance to therapy. Additionally, in cases where tumor tissue is not available, clinicians may rely on cell-free DNA (cfDNA) testing as a surrogate for comprehensive tumor testing. While some gene rearrangements are well detected, current commercial liquid biopsy assays exhibit low sensitivity for fibroblast growth factor receptor (*FGFR*) rearrangements. *FGFRs* are altered in ∼2.5% of all cancers, including *FGFR2* rearrangements in 10% of intrahepatic cholangiocarcinoma and *FGFR3* point mutations and rearrangements in 10-15% of urothelial carcinoma. Therefore, we developed and analytically validated FGFR-Dx, an *FGFR*-focused cfDNA assay with improved sensitivity for *FGFR* rearrangements. FGFR-Dx comprehensively targets the introns in *FGFR1-3* previously shown to be involved in gene fusions as well as all coding exons. Custom *FGFR* synthetic reference standards representing both single nucleotide variants (SNVs) and gene rearrangements were utilized at a range of variant frequencies and revealed a detection limit of 0.5% with sensitivities of 97.2% and 92.9% for SNVs and rearrangements, respectively. Furthermore, FGFR-Dx detected rearrangements and identified the intronic breakpoints from cfDNA collected from 13 of 15 patients with known FGFR fusions.

## INTRODUCTION

Liquid biopsy has emerged as an important tool for diagnostic testing in advanced cancer patients to detect predictive or prognostic biomarkers and clinically actionable genomic alterations. Hybridization capture-based multi-gene panels have been developed for liquid biopsy of cell-free DNA (cfDNA) to detect alterations in diverse oncogenes, such as *EGFR, BRAF, ALK*, and *RET*. More recently *FGFR* genomic fusions have become a therapeutic target in cholangiocarcinoma, bladder carcinoma, and 8p11 myeloproliferative neoplasms. Additionally, ongoing clinical trials of FGFR kinase inhibitors are demonstrating clinical benefit in diverse solid tumors, such as pancreatic and uterine cancers. While the sensitivity of all fusion detection is often extrapolated from *ALK* and *RET* fusions to other gene fusions, such as *FGFR*, recently the sensitivity of commercial liquid biopsy assays for *FGFR* gene fusions has proven to be low.

In a recent study of 1671 patients with biliary tract cancers, Berchuk *et al*. reported a cfDNA liquid biopsy sensitivity of only 18% for *FGFR* fusions, detecting *FGFR* fusions in only 12 of 67 patients with known *FGFR* fusions based on corresponding tumor tissue^1^. Importantly, this study demonstrated sufficient tumor fraction of cfDNA. We propose several inherent reasons that have led to reduced sensitivity for *FGFR* fusions in current approaches and assays. First, in practice, most analytic validations of multi-gene panels for liquid biopsy have focused on positive controls involving *ALK, RET, NTRK*, and/or *ROS1* gene fusions, but have not assessed detection of *FGFR* gene fusions. Therefore, these assays did not adequately validate detection of *FGFR* fusions^2-4^. There are relatively few *FGFR* gene fusion positive cell lines, and patient tumor tissues also have limited availability. Off-the-shelf cfDNA positive controls can be purchased but do not have *FGFR* fusions. Second, gene fusion breakpoints often occur in intronic regions, vary in size, have diverse gene partners and, as a result, may lead to variable capture and performance for hybridization-based capture panels. Routine probe designs typically omit repeat masked regions of introns due to multi-mapping and constraints for sequencing bandwidth for large multi-gene panels. Third, bioinformatics approaches for detecting gene fusions are complex and have many false positives making clinical assay development more challenging than detection of single nucleotide variants.

To address this unmet need for patients who have *FGFR* rearrangements, we present an analytic validation of an *FGFR*-focused assay for cfDNA liquid biopsy. To overcome the aforementioned challenges, we developed a set of 24 synthetic oligonucleotides as positive controls for gene rearrangements and single nucleotide variants (SNVs) and utilized a cohort of *FGFR* fusion-positive patient blood samples. Next, we developed a custom probe design targeting *FGFR1, FGFR2*, and *FGFR3* genes, including comprehensive capture of intron regions involved in rearrangements in addition to all coding exons. An accurate liquid biopsy assay for *FGFR* rearrangements can facilitate rapid molecular diagnosis in patients with cholangiocarcinoma, bladder cancer, and other solid tumors. Further, a small, focused assay is cost-effective for repeated serial testing and can be used to characterize the kinetics and clearance of *FGFR* rearrangements in patients receiving therapy.

## MATERIALS AND METHODS

### FGFR-Dx content and probe design

Biotinylated 120-mer probes (Integrated DNA Technologies, IDT™) were designed to target all exons and select introns of *FGFR1* (NM_015850) (introns 4, 5, 9, 14, 16, 17), *FGFR2* (NM_000141) (introns 4, 6, 9, 17), and *FGFR3* (NM_000142) (introns 5, 6, 11, 12, 13, 14, 16, 17). Probes were tiled with 60 base pair (bp) overlap. Probes were separated into low-risk and high-risk pools based on sequence specificity determined by both IDT’s internal assessment and the presence of more than one BLAT result with score >50. If either assessment suggested low specificity, the probe was placed in the high-risk pool.

### Custom FGFR synthetic reference materials

We generated 175 bp double stranded synthetic oligos to recapitulate the DNA fragment size observed in cfDNA extracted from plasma (gBlocks® Gene Fragments, IDT). A total of 48 synthetic oligos were designed, representing 12 *FGFR* fusions, 12 *FGFR* SNVs, and 24 matched wildtype controls. Individual oligos were diluted to 5 nM. A “mutant mix” (containing 50 pM each of 12 SNVs and 12 fusions) and a “WT mix” (containing 50 pM each of 24 matched wildtype controls) were generated. The “mutant mix” was then diluted 1:3 with the “WT mix” to generate the highest mutant variant allele frequency (VAF) mix of 25%, and 7 subsequent 1:1 serial dilutions were generated to cover a wide range of variant allele frequencies. We then spiked these synthetic sequence mixes into patient cfDNA at roughly equal molar concentration resulting in a further two-fold dilution and yielding final intended VAFs ranging from 12.5%-0.1%.

### Library preparation, hybridization-based capture and sequencing

10 ng plasma cfDNA (with or without synthetic mixes) or whole blood genomic DNA was used as input for library preparation with KAPA HyperPrep Kit (Roche) using dual-index adapters with a single unique molecular identifier (xGen UDI-UMI adapter, Integrated DNA Technologies (IDT™)). For hybridization and capture with our custom FGFR-Dx probes, 500 ng of library from 12 samples were pooled (6 μg total hybridization input) as previously described^5^. Sequencing was performed on an Illumina NovaSeq 6000 instrument.

### Unique molecular identifier (UMI)-informed analysis

UMI-aware demultiplexing from raw BCL/CBCL files was performed with Illumina’s software bcl2fastq version v2.20.0.422, running on Java version 1.8.0. Initial alignment to hg19 was performed with BWA version 0.17.17 using the “mem” algorithm, GATK version 4.0.10, and SAMtools v1.16.1^6-8^. Consensus calling was performed using fgbio tools version 2.1.0, requiring at least 3 reads per UMI family for SNVs and 1 read per UMI family for rearrangements, composite bases with quality <30 masked, 5% maximum no-calls, and maximum per-base error rate 0.1^9^. Consensus reads were re-aligned with BWA mem. SNVs were called utilizing VarDict version 1.8.2, with 0.1% minimum variant allele frequency, minimum base quality of 20, and minimum mapping quality of 10.^10^ Fusions were called with Manta version 1.4.0 and fusion supporting reads were combined if multiple breakpoints were called within 150 bases of each other.^11^ Computations were performed on the Owens cluster at the Ohio Supercomputer Center (https://www.osc.edu/).

### Sequencing coverage and computational down-sampling

To identify the number of input fragments required to yield a desired coverage of ∼2000x for both SNVs (using min3) and rearrangements (using min1), we stochastically down-sampled input fragments and assessed coverage. We determined that approximately 3.0 × 10^6^ and 0.5 × 10^6^ input fragments, for SNVs and fusions respectively, are required to yield ∼2000x coverage. One replicate for Mix 8 was excluded from SNVs analysis (min3) due to poor coverage.

### Calculating expected VAFs from mixtures

Since the observed max VAFs (Mix 1) varied from intended of 12.5% VAF, we determined expected variant allele frequencies for individual variants in each mix by applying a simple linear regression using the VAFs for each variant from Mixes 1-3 (for SNVs) or 1-3 (for rearrangements). Slopes were then multiplied by actual VAFs to calculate an expected VAF for each variant in each mix. Linear regression of expected vs. observed VAFs resulted in coefficients of determination (R^2^) >0.94 for each variant.

### Patient sample collection and isolation of DNA

Patients were consented to genomic testing under the institutional review board-approved study OSU-13053, *Precision Cancer Medicine for Advanced Cancer through High-throughput Sequencing*. This study allows for serial evaluation of blood and tissue specimens for genomic analyses (NCT02090530). Whole blood samples from patients were collected in EDTA or cfDNA BCT® (Streck) tubes and processed within 6 hours or 7 days of collection, respectively. Plasma was isolated by centrifugation at 1,000 rpm for 10 minutes and stored at -80°C. Genomic DNA was isolated from 400 μL whole blood using the QIAamp DNA Blood Mini Kit (QIAGEN). Prior to cfDNA isolation, plasma samples were spun at maximum speed (14,000 rpm) for 10 minutes. cfDNA was then isolated from 3-4 mL of supernatant using the QIAamp Circulating Nucleic Acid Kit (QIAGEN). Quality control was performed using Qubit, NanoDrop and TapeStation4150.

## Data availability

Sequencing data will be available upon request. Data will be deposited in dbGAP.

## RESULTS

### Probe design and generation of custom *FGFR* reference standards

Our custom *FGFR*-focused cfDNA assay (FGFR-Dx) detects SNVs and gene rearrangements in *FGFR1-3*. The probes for FGFR-Dx are divided into two pools: low-risk and high-risk. High-risk regions involved probes that were considered multi-mapped (**Methods**), which comprised 11% of the targeted region, and all others were considered low risk.

Current commercially available cfDNA reference standards include only one SNV in *FGFR3* (S249C, Seraseq® and ZeptoMetrix®) and no gene rearrangements involving any *FGFR* genes. Therefore, robust analytic validation of FGFR-Dx required the generation of custom reference standards. We designed 175bp synthetic oligonucleotides representing 12 *FGFR* SNVs and 12 *FGFR* gene rearrangements (“mutants”, four each for *FGFR1-3*). Additionally, we designed oligos representing the corresponding wild type sequences for these same 24 genomic positions (48 total synthetics). Due to the repetitive nature of high-risk regions in *FGFR* introns, it was not feasible to design synthetic oligos in these regions. Therefore, all synthetic oligos used for this validation were present in low-risk regions. Additionally, our initial validation did not include small insertions/deletions (indels) or copy number variations (CNVs).

We generated 8 serial dilutions using synthetics (Mixes 1-8) with intended variant allele frequencies (VAFs) ranging from 12.5%-0.1% (**Methods, Figure 1A**). Each synthetic mix of was spiked into 10 ng of the same patient cfDNA. In contrast to other commercial cfDNA assays which use a range of 10, 20, and 50 ng of DNA input for library generation, we focused our validation on 10 ng input since 20 or 50 ng is not always feasible from patient samples. Equimolar mixtures of synthetics and cfDNA were generated and subsequently underwent library preparation, hybridization-based capture with low-risk probes, and Illumina sequencing. Two technicians independently prepared 24 libraries for sequencing (**Figure 1B**).

**Figure 1.**
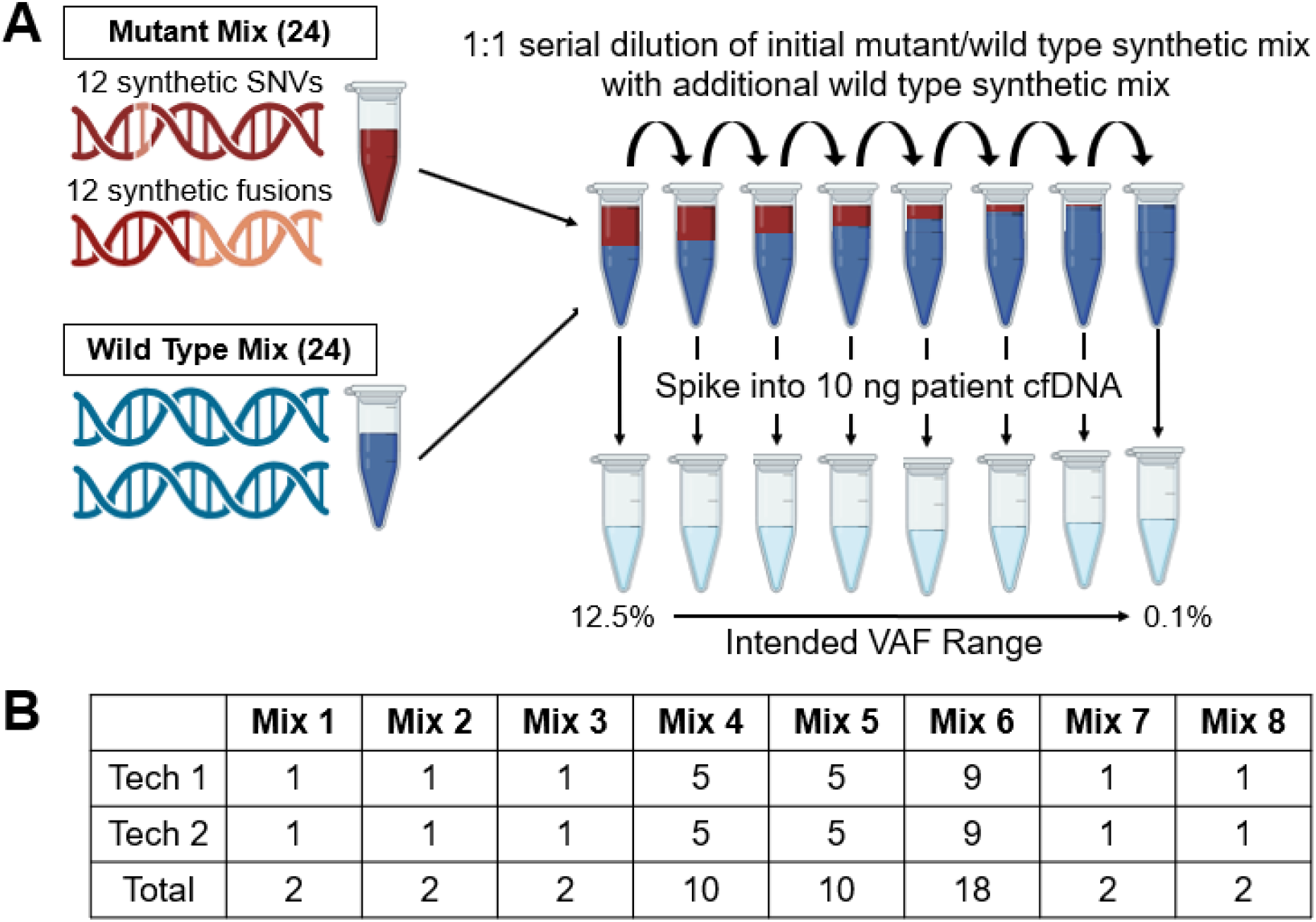
Validation strategy. **A**. Schematic of synthetic mixing strategy and serial dilutions to generate 8 mixes. Each of the 8 mixes was spiked into background cfDNA at an equimolar concentration and underwent library preparation, hybridization/capture using custom FGFR probes, and sequencing. **B**. Summary of independent samples prepared by each technician.

### Coverage of synthetic regions and down-sampling

Validation samples were multiplexed with the goal of generating approximately 2.20 × 10^7^ raw fragments per sample. We achieved an average of 2.02 × 10^7^ raw fragments per sample, with a minimum of 1.45 × 10^7^ and maximum of 2.56 × 10^7^. We utilized a two-tiered analysis approach, analyzing samples using both a minimum read family size of three (min3) and one (min1) for detection of SNVs and gene rearrangements, respectively. Despite limited utility of consensus calling for rearrangements, UMI-informed analysis with min1 rescues a significant portion of duplicate fragments with unique UMIs. This led to an average coverage of ∼4,500x for min3 and ∼7,000x for min1. With the addition of synthetics, the average coverage across these 175 bp regions increased to ∼7,500x (min 3) and ∼11,200x (min 1). Based on previously published cfDNA assay validations and our own retrospective data from patient cfDNA samples, this level of coverage is not necessary for robust variant detection and would be costly for prospective patient samples^2-4,12^. Therefore, we computationally down-sampled input fragments to achieve coverage of ∼2,000x for both min3 and min1 analyses (**Figure 2A, 2B**). All subsequent analyses were performed on this down-sampled data.

**Figure 2.**
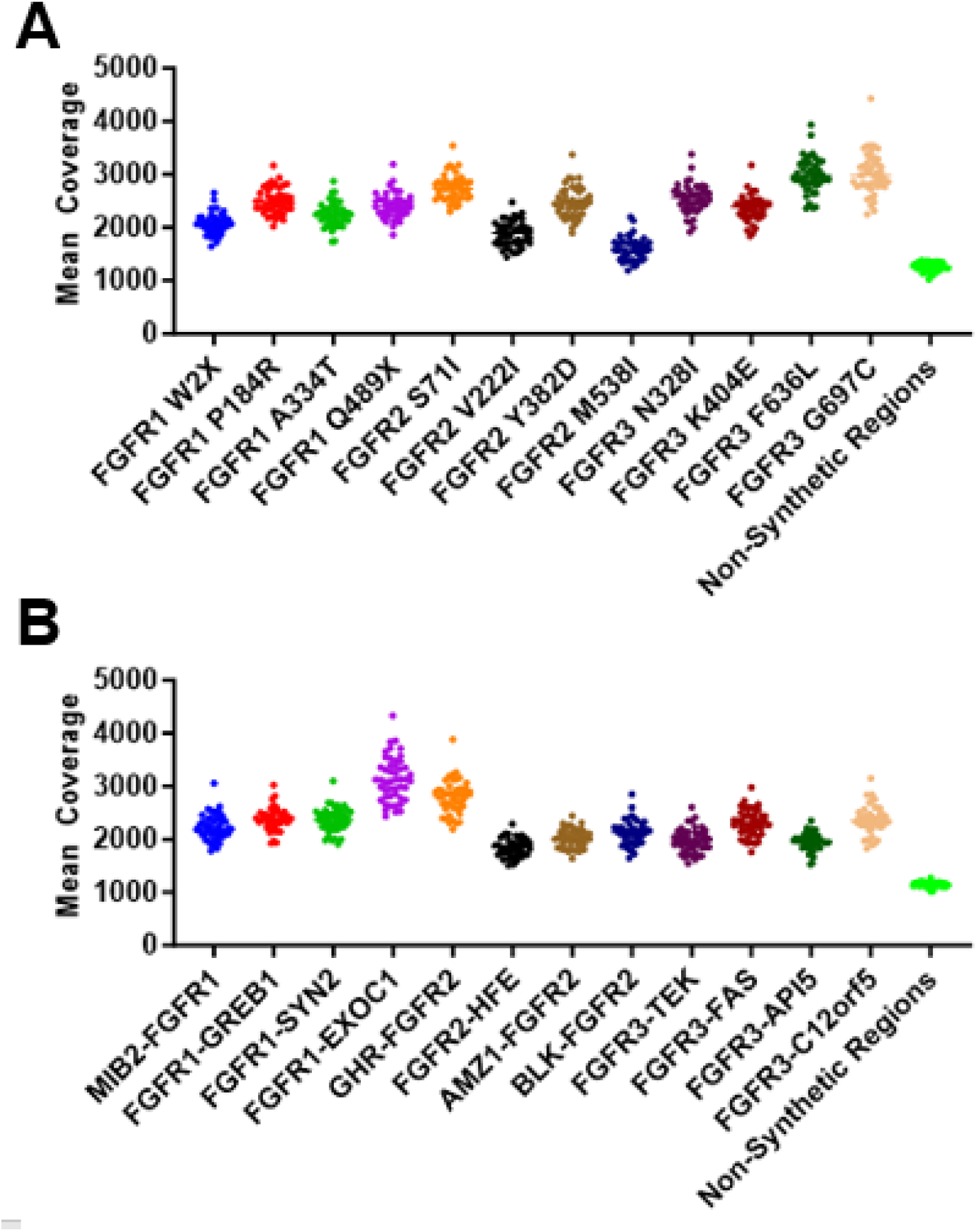
Down-sampled coverage in synthetic spike-in regions. **A**. Mean coverage of each 175bp synthetic SNV region after processing with 3 × 10^6^ input fragments using UMI minimum read family size of 3. The mean coverage of the remaining portion of the targeted region is illustrated last in lime green. n = 47 for each region. **B**. Mean coverage of the 87bp FGFR portion of each synthetic rearrangement region after processing with 5 × 10^5^ input fragments using UMI minimum read family size of 1. The mean coverage of the remaining portion of the targeted region is illustrated last in lime green. n = 48 for each region.

### FGFR-Dx accuracy

To assess sensitivity of FGFR-Dx, a total of 48 samples of plasma cfDNA with synthetic mixes were sequenced (**Figure 1B)**. Each sample contained 12 SNVs and 12 rearrangements, for a total of 576 possible events across a range of VAFs for each mutation type. Although we made 8 serial dilutions with intended VAFs from 12.5%-0.1% for each individual variant across the mixes, we observed some variability in the actual VAFs called for each variant. Therefore, we calculated expected VAFs empirically (**Methods**). Based on these expected VAFs, we determined assay sensitivity independently for SNVs and rearrangements across a range of detection limits and alternate read counts (0.1%-1.0%, 1-10, respectively) (**Figure 3A, 3C)**. Additionally, we calculated the positive predictive value (PPV) across these same detection limits and alternate read counts (**Figure 3A, 3C**). At a limit of detection of 0.5%, FGFR-Dx was 97.2% sensitive for SNVs and 92.9% sensitive for rearrangements with alternate read counts of 6 and 8, respectively. The PPV using these same parameters was 99.1% for SNVs and 99.3% for rearrangements. To ensure our validation cohort included representation near our detection limit, we plotted the observed VAFs for all true positive SNVs and rearrangements (**Figure 3B, 3D**). Approximately 17% of variants were detected at or below 0.5% for both SNVs and rearrangements (n=47 and n=29, respectively). Further, close to half of variants were detected at or below 1% VAF (42.3% for SNVs (n=116) and 51.5% for rearrangements (n=87)).

**Figure 3.**
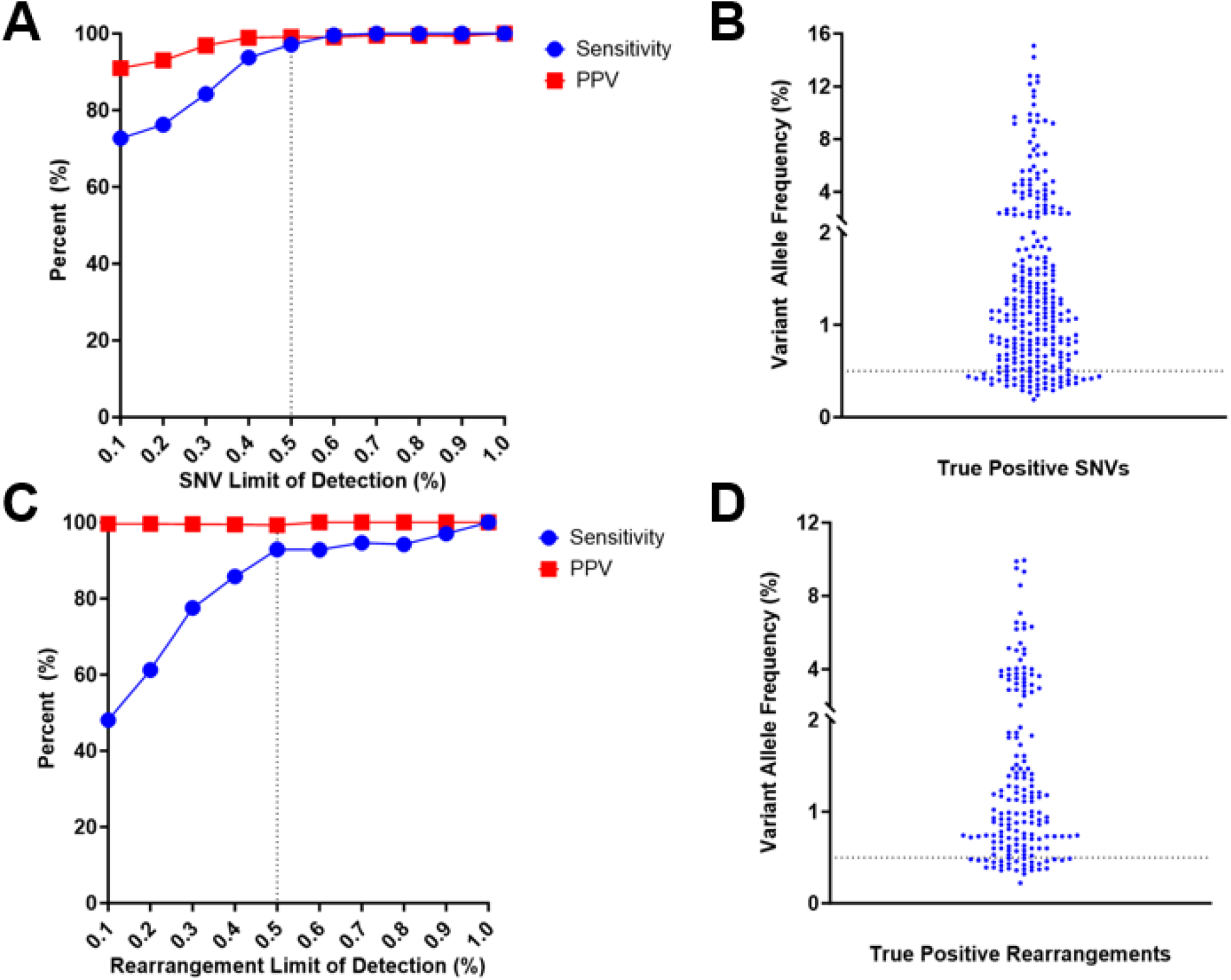
Sensitivity and PPV for SNVs and rearrangements. **A**. Sensitivity and PPV of FGFR-Dx for detection of SNVs across indicated detection limits with alternate read count ≥6. **B**. VAFs of true positive SNVs observed at detection limit of 0.5% with alternate read count ≥6 (n=274). **C**. Sensitivity and PPV of FGFR-Dx for detection of rearrangements across indicated detection limits with alternate read count ≥8. **D**. VAFs of true positive rearrangements observed at detection limits of 0.5% with alternate read count ≥8 (n=169).

### FGFR-Dx precision

To assess the reproducibility of FGFR-Dx, two different technicians prepared libraries from the same starting mixes (duplicates from Mixes 1-8). We aimed to examine reproducibility at the established detection limit of FGFR-Dx (0.5%). Therefore, our analysis focused on variants that were expected to be detected at a VAF of 0.5% or higher. Using these parameters, reproducibility for FGFR-Dx was 99% for SNVs and 97.9% for rearrangements.

To assess assay repeatability, the same technician prepared libraries from Mix 4, 5, and 6 in quadruplicate. Like reproducibility, we focused our assessment of repeatability on variants expected at or above our established detection limit of 0.5%. This led to a repeatability of 97% for SNVs and 88.6% for rearrangements.

### FGFR-Dx detects rearrangements in patient cfDNA

To examine the performance of FGFR-Dx in patient-derived samples, we applied the assay on 15 cfDNA samples collected from patients with tissue confirmed *FGFR*-altered cancers. For patients with gene rearrangements, detailed information regarding the genomic positions of breakpoints were not provided in most cases (only gene level partners involved). Therefore, initial samples from all patients were run with high and low-risk probes to ensure comprehensive assessment and the greatest probability of detecting rearrangements. An example of sequencing coverage in intron 17 of *FGFR2* for a patient with only low-risk probes vs. high + low-risk probes is illustrated in **Figure 4A**. We demonstrated that the addition of high-risk probes rescues coverage of the multi-mapped regions of *FGFR* genes (**Figure 4B**). For sequencing efficiency, if patient cfDNA breakpoints were determined to be in a low-risk region, all subsequent samples were captured using only the low-risk probes.

**Figure 4.**
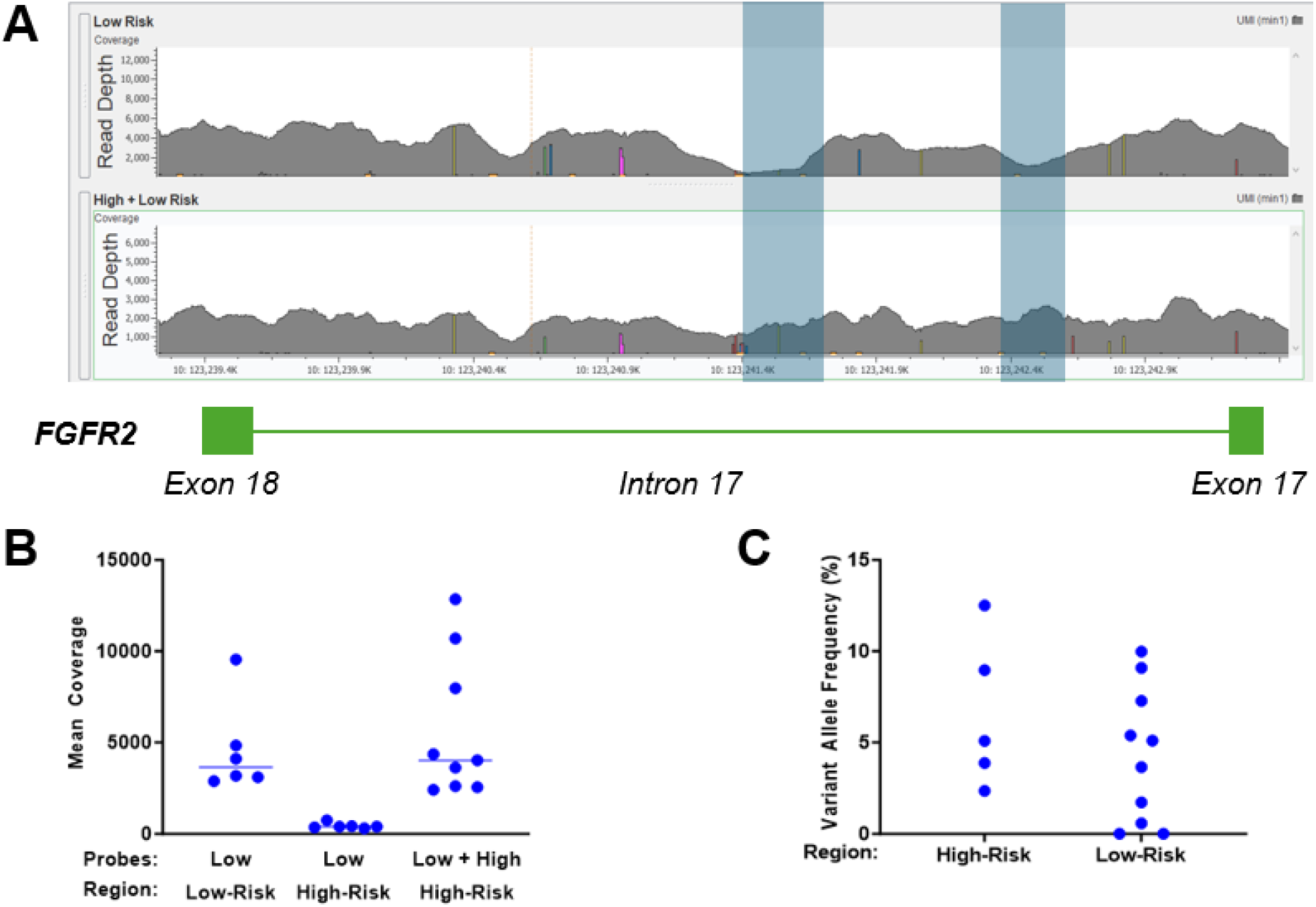
Detection of FGFR breakpoints in high-risk regions. **A**. GenomeBrowse screenshot depicting read depth in intron 17 of *FGFR2* for a patient sample captured with low-risk probes only (top) and low + high-risk probes (bottom). Blue highlights indicate high-risk regions. **B**. Mean coverage for either low or high-risk portion of targeted capture region in samples captured with indicated probes. Horizontal lines represent median. **C**. FGFR-Dx VAFs of FGFR rearrangements from 15 patient cfDNA samples with breakpoints located in high-risk (n=5) and low-risk (n=10) regions.

In a cohort of 15 patients, including 5 with high-risk breakpoints, FGFR-Dx detected rearrangements in 13 of 15 (86.7%) patient cfDNA samples. There was no substantial difference in calculated VAFs between high-risk and low-risk regions (**Figure 4C**). For the two instances where rearrangements were not detected, FGFR-Dx did detect the expected rearrangement from the corresponding tumor tissue sample (both breakpoints in low-risk regions), suggesting that low tumor burden or minimal shedding of cfDNA may explain the lack of detection in plasma. Consistent with this premise, both patients had small, single lesions observed on imaging at the time of blood collection.

## DISCUSSION

Through a novel probe design for improved coverage of *FGFR1-3* and the use of diverse positive controls for fusions, we have established a highly accurate and precise assay, FGFR-Dx, to detect *FGFR* fusions and mutations. The analytic validation was completed using both synthetics and patient blood samples, demonstrating accurate detection of fusions at a 0.5% VAF limit of detection, which is comparable or lower than commercial assays (0.5% to 1%). Furthermore, through a small, inexpensive panel focused on just *FGFR* genes and not 100+ genes, the cost of the assay affords an opportunity to test patients more frequently compared to other cfDNA studies. While this assay was developed based on an unmet need and observations that existing assays were not effective in cholangiocarcinoma, it is now evident that this assay is applicable to multiple solid tumors with *FGFR* fusions^13-16^.

Detection of diverse *FGFR* fusions requires a comprehensive capture of all introns involved in known breakpoints. While *FGFR* fusion partner genes are diverse, the breakpoints for *FGFR2* and *FGFR3* most commonly occur within the final intron. Through evaluation of the canonical introns where *FGFR* fusions occur, we identified approximately 15% of the targeted introns (50,471bp) as being highly repetitive regions that are omitted from standard probe designs by RepeatMasker and/or proprietary design preferences. The inclusion of these highly repetitive regions results in probes with multiple genomic matches and increases the rate of capturing off-target genomic regions. While this leads to a loss of sequencing efficiency, by focusing on *FGFR* genes the assay is still capable of achieving sufficient on-target capture and coverage for UMI-based analysis at a reasonable cost. We observed 5 patients with *FGFR* fusion breakpoints within these highly repetitive regions and predictably not detected by commercial assays, demonstrating the value of expanded capture of introns. However, it is unclear how many patients have fusions in these repetitive regions, or how many fusions are still being missed by current assays. Without using positive controls for FGFR rearrangements in the validation of larger multi-gene panels, we speculate that cut-offs for *FGFR* may be more stringent for the sake of retaining specificity at the cost of losing sensitivity.

Synthetic controls can overcome the challenge of commercially available controls lacking *FGFR* fusions and of limited cell lines available with *FGFR* fusions. Nearly 100% of liquid biopsy analytic validations rely on use of cfDNA mixtures as positive controls that span point mutations, copy number alterations, and rearrangements^2-4^. Such commercially available reagents use either cell lines, engineered cell lines, or synthetic DNA oligos to build their positive controls. For fusions, these commercially available reagents include *ALK, RET, ROS1*, and *NTRK* fusions, but do not include *FGFR* fusions. Thus, synthetic oligos for *FGFR* fusions can serve to represent these underrepresented and under evaluated oncogenic genomic alterations. Synthetics or contrived mixtures do not replace the examination of real-world blood samples but can be complementary to analytic validation or quality control evaluations of liquid biopsy assays.

The most common clinical application for liquid biopsy is for molecular classification and identification of driver oncogenes that can guide selection of a targeted therapy. There is great interest and unmet need for application of liquid biopsy in intrahepatic cholangiocarcinoma where *FGFR* fusions occur in 10% of patients. For cholangiocarcinoma, tumor tissue is not always readily available for molecular testing. In a substantial portion of patients, a liver biopsy demonstrates poorly differentiated carcinoma requiring many sections for immunohistochemistry in the workup, and cholangiocarcinoma is often the pathologic diagnosis of exclusion. Further, the location of cholangiocarcinoma in the liver is often not feasible for core needle biopsy and may only yield a fine needle biopsy. Additionally, up to 10-20% of some tumor biopsy specimens may have insufficient quality for sequencing. However, a recent study raises concerns that current liquid biopsies lack the sensitivity to detect *FGFR* rearrangements. Berchuk *et al*. applied Guardant360 to biliary tract cancers and identified 12 fusions in cfDNA out of 67 patients with known *FGFR* fusions based on corresponding tumor tissue^1^. Other liquid biopsies have not reported on their sensitivity for *FGFR* fusions, and instead extrapolate sensitivity for all fusions based on assessment of *ALK, RET, ROS1*, and/or *NTRK* fusions. A small pilot study reported on feasibility of molecular diagnosis from cfDNA in newly diagnosed patients with cholangiocarcinoma or pancreatic cancer^17^. However, it is important to know whether the assay is clinically validated to detect *FGFR* fusions.

A second application of *FGFR* liquid biopsy is determination of acquired mechanisms of resistance. Wu et al, reported a comprehensive summary of acquired mutations in *FGFR* kinase domain that impart resistance to reversibly binding FGFR kinase inhibitors^18^. They reported that 49 of 82 patients harbored at least one mutation in the *FGFR2* kinase domain, most commonly N550 and V565 amino acids. As second generation FGFR kinase inhibitors such as futibatinib (recently FDA-approved in cholangiocarcinoma) and other investigational agents in clinical trials, clinical assessment of emerging resistance mutations can help guide the role and selection of subsequent therapies for patients. While FGFR-Dx has improved sensitivity for fusions and can also detect point mutations, the focus on *FGFR* genes has introduced some limitations. While secondary *FGFR* mutations occur in the majority of patients (∼60%), some patients instead have acquired genomic alterations in MAP kinase and PI3 kinase pathway genes such as *BRAF* and *PIK3CA*. Similarly, not all patients with cholangiocarcinoma have *FGFR* driver mutations, and may harbor other genomic alterations involving *IDH1, ERBB2*, and others^19^. Further, our initial design focused on *FGFR1*-3, and did not include *FGFR4* since there are exceptionally rare alterations in this gene. While this is a limitation of an *FGFR*-focused assay, this can be addressed through expanding the panel to include additional genes of interest including *FGFR4, IDH1/2*, and *ERBB2*.

Strengths of FGFR-Dx, a smaller panel focused on fewer genes, include not only improved sensitivity for *FGFR* fusions, but also new opportunities to apply this small, cost-effective assay serially over time and the course of a patient’s treatments. Thus far, liquid biopsy studies for oncogene-addicted cancers such as *EGFR* and *KIT*, have evaluated few patients at timepoints corresponding to pre-treatment and CT scans performed every 2-3 months^20,21^. The disadvantage of this approach is that it misses an opportunity to evaluate early kinetics of cfDNA in response to treatment that may be clinically informative. Liquid biopsy results have the potential to guide clinical decision making in advance of CT scans, thus enabling earlier interventions before metastatic disease has advanced too far. Studying the kinetics of *FGFR* cfDNA during therapy can unlock the potential for assessing response to therapy, which can help clinicians choose alternate therapies, adjust doses, and plan for future therapies or clinical trials.

Beyond *FGFR* fusions, our approach has broader implications for improving the detection of other gene fusions via liquid biopsy assays. NTRK kinase inhibitors are now approved for any tumor with fusions involving *NTRK1, NTRK2*, and *NTRK3*. Notably, *NTRK* fusions occur in canonical introns and are massive compared to introns for *FGFR* genes, making capture and detection of *NTRK* fusions even more challenging. In a recent study, cfDNA testing detected *NTRK* gene fusions in only 33% (33/99) of patients with known *NTRK* fusions based on tumor tissue testing^22^. In another example, Supplee *et al*. sought to improve cfDNA detection of *ALK* fusions in lung cancer^23^. The authors proposed that shorter probes (∼40 nucleotides) may improve capture of these regions and thereby improve detection of *ALK* fusions. They were able to detect 13 of 16 fusions compared to another assay (7 of 16). In a recent pan-cancer study, *FGFR* fusions were seen in ∼5% of cholangiocarcinoma, compared to the expected prevalence of ∼10%^24^. These studies underscore a critical gap between analytic validations that have extrapolated accuracy for many fusions. Real-world clinical validation of rearrangements in patients, provide an opportunity to improve rearrangement detection from cfDNA.

In summary, liquid biopsy has emerged as an important clinical tool for making a molecular diagnosis in patients with advanced cancer, and FGFR-Dx represents an important advance for improving accuracy for detecting *FGFR* as well as other kinase fusions. Our approach outlines how synthetics and enhanced designs for capture can be combined to improve accuracy. We propose that frequent monitoring of fusions during therapy can facilitate clinical applications to guide therapeutic decision making.

## Data Availability

All data produced in the present study are available upon reasonable request to the authors

## ACKNOWLEDGEMENTS

We are grateful for patients who have participated in clinical research studies that enabled this work. We express our gratitude for the Gateway for Cancer Research, Pelotonia, and the Comprehensive Cancer Center at The Ohio State University. We are thankful for the Ohio Supercomputer Center for providing the computational space on which this work was done. This work was supported by NCI UH2CA262220 grant. A.P. and R.B. were supported by Pelotonia fellowships. L.S. and R.B. were supported by T32 fellowships.

---

**Table 1.** FGFR-Dx Precision.

| **For expected**<br>**VAFs≥0.5%** | **Reproducibility**<br>**(Mixes 1-5)** | **Repeatability**<br>**(Mixes 4 + 5)** |
| --- | --- | --- |
| **SNVs** | **99% (113/114)** | **97% (163/168)** |
| **Rearrangements** | **97.9% (92/94)** | **88.6% (78/88)** |
| **Table 1. FGFR-Dx Precision.** |  |  |

